# Health-system readiness for integrating ceiling nets into malaria vector control in Kenya: perspectives of health managers

**DOI:** 10.64898/2026.07.02.26357099

**Authors:** Samuel M. Mbugua, James Kongere, Wataru Kagaya, Peter Mwaura, Yura Ko, Protus Omondi, Milton Mujuni, Anna Kasililika, Isaac Kisiangani, Bernard N. Kanoi, Akira Kaneko, Jesse Gitaka

## Abstract

Malaria transmission has been reduced considerably in some Kenyan regions; however, it remains high in the Lake Victoria basin. Ceiling nets fall under house screening as interventions in vector control under the World Health Organization’s Guidelines on Malaria and has a conditional recommendation with low-to moderate-certainty evidence of impact against malaria. This study aimed at assessing the perceptions of health system managers on the health system readiness in integrating ceiling nets into malaria vector control in and implementation pathways.

A cross-sectional, qualitative, action research design was used aimed at providing actionable solutions to health systems challenges in the inclusion of ceiling nets as a complementary vector control strategy in the comprehensive approach on malaria prevention in Suba West sub-county, Homa Bay county. Key-informant interviews on perspectives on health system readiness for ceiling net integration into routine programmatic vector control strategies were conducted among health system managers. Qualitative data was captured using audio tapes and field notes, transcribed and analyzed using case-based in-depth inductive and deductive analysis using a thematic framework to classify and organize data into themes.

Key emergent themes included perception and experiences in malaria vector control strategies, role of community health in malaria vector control strategies, systemic barriers and facilitators in malaria vector control, perspectives on integration of the ceiling nets in malaria vector control. This study demonstrates that health system managers perceive ceiling nets as a feasible, acceptable, and operationally integrable complement to existing malaria vector control strategies, while also identifying concrete system bottlenecks that must be addressed for scale-up. The ceiling nets showed high acceptability, appropriateness of fit in structurally heterogenous housing structures and high feasibility as an addition to a comprehensive complementary vector control strategy.

The findings inform decision-ready implementation pathways of complementary ceiling nets in existing vector control strategies. Ceiling nets are perceived by frontline health system managers as a system-ready, acceptable house-modification intervention that can close critical protection gaps left by bed-net–centric malaria control, provided community health platforms and supply chains are deliberately leveraged. Successful scale up implementation hinges on a structured approach of integration that includes community buy-in, task-shifting of community health promoters in ceiling net installation, effective supply chains, and robust monitoring and evaluation systems to track, maintain and assess system level indicators and community level indicators.

## Introduction

The current malaria control strategies in most regions in Kenya include distribution of insecticide-treated bed nets, improved malaria case management, and targeted vector control strategies such as indoor residual spraying and seasonal malaria chemo prevention. Despite the implementation of the Kenya’s comprehensive Approach on Malaria Prevention [1], the disease remains a persistent threat to the country’s public health system. The cornerstone of malaria elimination efforts rests in vector control strategies owing to facts such as emergence of drug resistance, unaffordable potent antimalarials [2], and challenges in the malaria vaccination programming related to vaccine’s complex four-dose schedule, health system barriers and logistical constraints [3]. The optimization of robust vector control strategies is highly dependent on entomological surveillance crucial for malaria elimination success ([4]. These efforts have not been without key challenges such as the effect of climate change on modifying quality and efficacy of quality vector control interventions [5]).

The most potent and important malaria vectors in Africa are readily vulnerable to population control, or even elimination, with long-lasting insecticidal nets (LLINs) because they predominantly feed on humans at times of the night when they are asleep inside houses [6]. The important limitation for the effective use of bed nets is that the nets are effective only when people are sleeping inside them [7]. While malaria transmission has been reduced considerably in some Kenyan regions, it remains high among the residents in the Lake Victoria basin. Intense malaria transmission is often observed in villages near lakes and large reservoirs in Africa [8].

Ceiling nets can benefit every member of the household since its use is not affected by similar factors that affect bedside nets. In a study on the effectiveness of screened ceilings over current best practice in reduction of malaria prevalence in Western Kenya, Minakawa et al. [8] indicated the effectiveness of ceiling nets in reducing indoor resting vectors in children aged between 7 months and 10 years old. A study evaluating the effectiveness of installing ceiling-mounted mosquito nets to reduce Anopheles mosquito density indoors and peri-domestic provided compelling evidence of significant reduction in exposure in a high-transmission structurally vulnerable community [9]. Ceiling nets fall under house screening as interventions in vector control under the World Health Organization’s Guidelines on Malaria [10]. House screening has a conditional recommendation with low-to moderate-certainty evidence of impact against malaria.

While studies evaluating the protective efficacy of ceiling nets are emerging [8], little is known about whether health systems are institutionally, operationally, and socially prepared to integrate them into routine malaria control, a prerequisite for policy adoption. This study was therefore conducted alongside a cluster-randomized controlled trial of Olyset® Plus ceiling nets in the Lake Victoria Basin, Kenya [11], which hypothesized that retrofitting Olyset® Plus LLINs into ceiling nets could complement existing malaria interventions and further reduce malaria transmission. The present study aimed to assess health system managers ‘perceptions of health system readiness for ceiling net integration to malaria control strategies, with a particular focus on service delivery within the health system.

## Methods

### Research design and setting

We employed a cross-sectional, qualitative, action research design aimed at providing actionable solutions to health systems challenges in the inclusion of ceiling nets as a complementary vector control strategy in the Comprehensive approach on malaria prevention in Mfangano island, Homa Bay county. The study obtained ethical clearance from Mount Kenya University’s Institutional Scientific Ethics review committee under the application approval number 1710. The key-informant interviews on perspectives on health system readiness for ceiling net integration into routine programmatic vector control strategies were administered to health managers in Mfangano island, Suba West sub-county. Purposive sampling was used to select the health managers drawn from the Sub-county Health Management Team in Suba Central. This is because they represented technical managers responsible for health service delivery in the sub-county, operational management, supervision and coordination. The recruitment strategy involved engagement of the health managers involved in implementation of malaria control strategies during the study sensitization and co-development workshops. Written Informed consent was sought from all participants. We interviewed a sub-county public health officer, a medical officer of health, sub-county community focal person, the Sub-county pharmacist, and a community health assistant. The highly specific participants provided insightful information into the inner setting health system readiness for integration of a complementary vector control strategy. Each interview was conducted for approximately one hour. The researchers acknowledged their positionality as an influence on the research process and employed reflexivity practices, including ran audit trail, consistent interview procedures, and iterative analysis, to minimize bias and ensure findings were grounded in participants’ perspectives. The study was nested within a five-year Interdisciplinary research for an integrated community-directed strategy for sustainable freedom from malaria project implemented in Homa Bay county.

### Implementation framework

We employed the Consolidated Framework for Implementation Research (CFIR) by Damschroder et al. [12] to gain understanding of variations in implementation success and provide recommendations for scaling up of ceiling nets in programmatic malaria vector control interventions. The model provided a domain-guided approach in the evaluation of health systems readiness for ceiling net integration emanating from the complexity and dynamic interplay of implementation processes, people, and health systems environment [13]. The CFIR constructs were instrumental in the development of the interview guide, and structural thematic analysis and coding. The themes were categorized based on domains of CFIR i.e. intervention characteristics of the ceiling nets, outer setting referring to external factors such as community needs and policies, Inner setting such as readiness and acceptance, characteristics of the end-users- cultural beliefs, knowledge and community champions of ceiling net use, and process factors referring to how the ceiling nets were rolled out and the roles of health system managers.

### Researcher reflexivity statement

This study was conducted and reported in accordance with the Consolidated Criteria for Reporting Qualitative Research (COREQ) guidelines [14]. The research team comprised public health researchers and implementation scientists with prior experience in malaria vector control and community health systems in Kenya. The researchers involved in qualitative data collection and analysis met at critical points of the data analysis to discuss emergent codes and themes. This was shared with the wider research team providing a combination of insights of those involved in data handling and members with a wider perspective of methodological and open disclosure matters. We acknowledge that our professional backgrounds may have influenced the interpretation of data toward health systems strengthening and scalability of the innovation. To mitigate potential bias, we maintained an audit trail of analytical decisions using project memos, used iterative coding, and held regular debriefing sessions to challenge assumptions. The lead qualitative researcher had no prior relationship with the participants, which helped maintain neutrality during interviews and analysis.

### Data collection

Qualitative data was captured using audio tapes and field notes, transcribed, and managed using QSR Nvivo 15 software. The key informant interview transcripts were coded and checked for consistency using a thematic framework to classify and organize data into themes using case-based in-depth inductive and deductive analysis. Using iteration as a component of action research, we developed a thematic framework which was updated following three rounds of iterative analysis. Thematic analysis charts were developed and categorized across the participants.

| Participant's role | Gender | Years of Experience |
| --- | --- | --- |
| Sub-county public health Officer (SCPHO) | Male | 8 years |
| Medical Officer of Health (MoH) | Male | 11 years |
| Community Health Assistant (HA) | Female | 17 years |
| Sub-County Pharmacist (SCP) | Male | 6 years |
| Sub-County Community Health Focal Person (CFP) | Female | 4 years |

## Results

### Theme 1- Inner setting: Perception and experiences in malaria vector control strategies

#### Sub-theme 1: Use of Long Lasting Insecticide treated Nets

Usage of the long lasting nets was perceived to be highly effective translating in low malaria endemicity in the communities. It is similarly cost effective due to the longevity nature of the nets and reduced need of purchasing other vector control methods like coils.

> “The long-lasting net has been very effective since introduction, and there is a low number of malaria cases compared to those areas that you never worked with. So, it’s very, very effective.” **KII-SCPHO**

> “Usage of the nets, is the best method of preventing the transmission of malaria in this community, and it becomes very difficult to explain to them that there are also other methods because they presume that the nets is what most of them know and is cost effective.” **KII-MoH**

In this malaria endemic region, the use of LLINs is complemented by other vector control strategies such as indoor residual spraying and malaria chemoprophylaxis. This indicates the multifaceted approaches employed by the health system in the prevention and management of malaria.

> “And from the county government and maybe national government as a whole, they offer… they give nets and anti-malarias are also given for free.” **KII-HA”**

Some of environmental vector control strategies that were highlighted focused on clearance of bushes and draining of stagnant water.

> **“**Ah, okay. Environmental vector control, the one we have, we always encourage people to clear around the surroundings. Then the stagnant waters which can be a breeding site for the malaria parasite, so we always encourage them that the stagnant waters they do away with. Clearing the bushes around them” **KII-CFP**

#### Sub-theme 2: Socio-cultural barriers to utilization of LLINs

The officials reported of socio-cultural barriers stemming from a lack of sensitization, bringing on the notion of potential side effects due to the usage of the LLINs.

> “So, you get some people believe that when subjected to that commodity, maybe they could give them other infections.” **KII-SCP**

It was also noted that there was improper use of LLINs as they would be used for other activities like fishing, drying silver cyprinid (omena) and fencing their garden.

> “Being that this area is a fishing area, some people are tempted to convert them to fishing nets, or use them to fence their kitchen garden. They also put it down there to make a rack where they dry their omenas (silver cyprinid). So, when they are torn, they presume that there is a second use. It’s like second-hand clothes which they can use for other purposes. But also, they are… there are situations where you can find the slightly… they look new nets converted, like when they want to sun dry their silver cyprinid or the small fish. They put it down the1010re, make a rack where they dry their omenas.**” KII-MoH**

Household sleeping arrangements proved to be a barrier to the utilization of LLINs, particularly due to the single net distribution per household, exposing some members to mosquitoes and malaria. The setup, especially in low socio-economic settings typically entails parents in a household sleeping on the only bed in the household and the children sleeping on the floor. This eventually results in reduced effectiveness of malaria prevention efforts as transmission remains high in children.

> “You see, within the department we presume that a family, like when we have mass net distribution, there is an assumption that one net should cover two people. Now, if you find maybe a father staying with the daughter in the same home, you will find that it is either the parent who will sleep under the net and the child will be out of the net. Also, you have a situation where people say that once you grow, you cannot sleep under the same roof with your mother or your son. Maybe they think it is a taboo. This might hinder the usage of that.” **KII-MoH**

Individual socio-behavioral barriers such as ignorance that resulted in inappropriate usage of LLINs were also mentioned as barriers to the effectiveness of LLIN utilization.

> **“**Okay, the barriers not too much. But most of the challenges that we can say is that some people don’t see the benefit. Like I can say that ah, not really from lack of education, but instead ignorance. At times when they’re distributed, you’ll find some people are using them to collect fish.”

> **“**Yes. For fishing. Some are also using them in other activities like covering the kitchen garden. Some we were able to find using them airing things like maize. So those are the only barriers. But using it, just like barrier.” **KII-CFP**

### Theme 2- Process: Community health in malaria vector control strategies

#### Sub-theme 1: Role of the community health promoters as facilitators of malaria vector control

Community health promoters are a crucial component in community malaria prevention and management and are perceived as gate keepers of the community. They sensitize, create demand and create awareness in the community.

> “CHPs are the key gatekeepers in the community. They are the people who will let the community know that this activity is taking place. They’re the people who can identify that a given household is using the net as required. So, in the activity, they’ll play a very major role if they are involved 100 percent” **KII-SCPHO**

Community health promoters comprise the community units and play an important role in ceiling net roll out requiring training in installation and demand creation for the ceiling nets. Their role as community gatekeepers positions them uniquely as the vital manpower needed in the installation of ceiling nets.

> “We have, community health assistants i.e. the CHAs manning each and every community unit, and these people are manning a given number of CHVs, and these CHVs are manning a given number of households. So, the people that are required for training are the community health assistants (CHAs), which in the island, we have six.” **KII-SCPHO**

> “The CHVs are the first level of management and if they do their work well, the workload in the facilities will be reduced and quite a good number of the community will be doing their own activities. First, the CHVs are involved in creating awareness within their villages. Currently we have around five community units out of eleven who have been trained on malaria case management and they are involved in testing their own community members and treating at the same time because they have been empowered.” **KII-MoH**

Their role in community advocacy in malaria is exemplified in the narration below which highlights several advocacy and health promotion roles.

> “The CHVs sensitize, and they also advise people to go test for malaria in the facilities. For the mothers they encourage them to maybe go for clinics so that they are advocated nets within the facility when they’re coming for their ANC visits. They are also tested for malaria just to see they’re negative. And if they have malaria interventions are taken… they’re taken through some interventions just to prevent and then… okay that is CHVs.” **KII-HA**

#### Sub-theme 2: Support for the community health promoters

The CHPs within the community health system remain an under-represented component of the health care workshop with lack of commensurate terms of service. CHPs are periodically supported by ongoing donor funded projects, however, the support is limited to the project’s duration and project activities. CHPs can be supported by enhancing the community health system through institutionalization to ensure reach and demand creation through provision of transport reimbursement, community dialogue forums and offering trainings to empower them.

> “To make sure that the CHVs reach everyone, you can support them by giving them transport reimbursement. You can as well support CHVs by organizing community dialogue.” **KII-SCPHO**

> “The empowerment is by training them, providing them with antimalarials and test kits followed by a monthly review of their activities so that at least the challenges that they face during that process are addressed.” **KII-MoH**

#### Sub-theme 3: Key decision makers in the community

The key community gate keepers and decision makers mentioned included CHPs, chiefs, assistant chiefs, village elders and religious leaders. Community engagement strategies include group empowerment forums (*chamas*) and funeral processions (*disco matanga*). Chamas (Swahili word for change) are self-organizing micro-saving groups that continue to be popular across Kenya particularly among women. The disco matanga is a traditional, often nighttime, event held to mourn and celebrate a life, sometimes used to help fund funeral expenses through social gatherings.

> “The CHVs themselves, administration, the chiefs, assistant chiefs, village elders, and even the religious leaders in some areas, and even the household heads, play a key role in making decisions.” **KII-SCPHO**

> “The chief barazas have been involved and of late even the fishermen out there, their *chamas*, have been involved, and also when you have something called *disco matanga* around. So, in that forum when you are given CHV or other professional who is on there like the Public Health Officer, they can get on that forum to give the information, this is the right way to go.” **KII-SCP**

The church in most rural facilities is an important component of community advocacy. The influence of religious leaders on socio-behavioral aspects of communities is immeasurable in guiding the advocacy efforts around acceptance of ceiling nets. Their inclusion in the study inception was a valuable resource in getting community buy in and eventual acceptance of the ceiling net.

> **“**They went out before the inception. Okay, I remember the inception meeting. They called different stakeholders. So, the stakeholders included people from the church, non-governmental organizations, and the government.” **KII-HA**

#### Sub-theme 3: Training of community resource persons on installation of the ceiling nets

The public health office indicated that they have capacity for coordination of training and installation of ceiling nets based on prior experiential work on vector control. The ministry of health supports ongoing activities in vector control.

> “I believe that, my office is very capable because this is not the first time that we are doing it, we’ve been doing mass net distribution all over the island, being supported by the Ministry of Health. We know where to store it and who to involve when doing this.” **KII-SCPHO**

Training of CHAs is vital in the monitoring of household utilization of ceiling nets.

> “The administration needs to be part and parcel of this. These six community health assistants need to be trained so that they can do close monitoring.” **KII-SCPHO**

### Theme 3- Outer setting: Systemic determinants of readiness in malaria vector control Sub-theme 1: Collaborative approach in the research process

The health managers indicated that there was use of an inclusive approach in the ceiling net roll out. It was however noted that they should be involved more especially at study conception and the implementation process to ensure security and safety of the ceiling nets upon storage. The lack of inclusiveness in project implementation has in the past resulted in disjointedness of the multi-stakeholder malaria control efforts with some of the health managers highlighting this as mentioned in the second quote below.

> “If any case our department will be given an opportunity; all the key stakeholders will be involved. Where to store them, security will be guaranteed.” **KII-SCPHO**

> **“**Two, what we will require so that the whole island can be, eh, supported, the… the people that you are working with, those who are installing the net, as a… as a department. Even if you ask me today who and who that you used the other time, I cannot even know.” **KII-SCPHO**

#### Sub-theme 2: Determinants of uptake and utilization of the ceiling nets

The identification of knowledge gaps and interventional needs is a valuable resource in strengthening of malaria preventive, control and elimination efforts (Rahim et al., 2024). Community Knowledge transfer to community members is a critical determinant of uptake and use of ceiling nets. There was emphasis on CHPs knowledge brokers in households and villages.

> “Once the household has gotten the right information and education on the best practices on how to use net, how to dispose it off and how to clean it when it is dirty, automatically they’ll use the ceiling net promptly. Giving the right information to the CHVs so that they can disseminate the same to the household donors, they’ll actually support it.” **KII-SCPHO**

Continuous awareness creation is vital to successful integration of ceiling nets into programmatic vector control strategies and adoption at the community level.

> “We also have to do proper awareness, give information fully and let everyone be involved. If we don’t give much information on why we are going to give that, we are not going to achieve much because some also can say, now we are using the ceiling one, what about the old bed nets we were using before?” **KII-SCP**

> **“**The only support which I know and we always use is just sensitization and we mobilize. Because when we mobilize the community, they always understand the benefit of the bed nets. So, when we tell them that this is the same but the makeup is different, they will also… always buy.” **KII-CFP**

The source of sensitization and awareness is critical towards determining the utilization of nets.

> “What is influencing the use of the ceiling nets, number one is the primary information given on what the net is going to do. During implementation, the primary information to be passed on is what we intend to achieve with ceiling nets, and we will have easy implementation and also the usage.” **KII-MoH**

The type of structural design is also a vital determinant of the efficacy and utilization of ceiling nets providing an insight to the behavioral adoption dynamics with the health managers indicating that the ceiling net was appropriate for traditional rural houses with thatched roofs or iron sheets and mud walls.

> “The structure has got a lot to do with the efficacy of the ceiling nets and installation. We find that, for example, in most areas, people construct the *mabati* houses. If they are flat, it becomes very difficult to install, or if every place is covered and you need to change that design, you must get this person convinced that that the only reason why I’m putting this is to protect you from getting the mosquito bites and getting the malaria there. The nature of the structures and how they are designed, will also affect the use of the ceiling nets.” **KII-MoH**

#### Sub-theme 3: Health systems factors affecting vector control programs in malaria

The office of the sub-county public health officer managed the distribution of LLINs primarily through various service delivery points i.e. antenatal clinics, household visits and mass net distribution campaigns.

> **“**Your bed nets, we have two types. For the pregnant mothers, the ones they are being given when they are pregnant during ANC. After they have been given, we must during household visitation, give a health talk on how to use it, the best way how to handle it and the best way to dispose it off if any case it is old or torn. So, for mass distribution ones, we also have to look into.” **KII-SCPHO**

There was indication of commodity quantification systems in place to ensure constant availability of LLINs by the sub-county pharmacist.

> “I normally monitor everything I chair the progress meetings and also have a Whatsapp Group with all of them. So, what I do in that is that I’ll check and know, what have you received, what do you have, what don’t you have, what is expiring soon so that we can do redistribution. We have a central store, and in this central store, we have to have all the tools which can help us receive it first. After that, we have to map how many facilities we have. If we are going to roll it out in the entire health facility around, because the facility also come with the region, we just check what’s the population there to help us quantify what they can use.” **KII-SCP**

> “Once you have the commodity, you have to monitor, how these people are using this particular commodity and in case there are some problems, you need to rectify as soon as possible.” **KII-SCPHO**

#### Sub-theme 4: Availability of collection tools for M&E of ceiling nets as a vector control intervention

There is need for development of M&E indicators tools to monitor the utilization of ceiling nets in the community providing vital data into routine programming of malaria preventive strategies.

> “We have some supporting tools which help us in the maternal unit, when we do the MCHH, but when it comes to the general population, we have to come up with a tool. We need to have a monitoring tool which will just help us to monitor who and which homestead, because we also have to check like the data coming from all the homestead.” **KII-SCP**

### Theme 4- Intervention characteristics: Perspectives on integration of the ceiling nets in malaria vector control

#### Sub-theme 1: Appropriateness of fit, acceptability, and feasibility of the ceiling net

Health managers perceived a reduction in malaria burden in households or clusters where ceiling nets had been implemented, although this qualitative observation requires triangulation with surveillance and trial data.

> **“**With support of the testing, the routine testing and the use of the ceiling nets, we can see that there is a bit, not even a bit, but there’s a very bigger difference in prevalence as compared to the previous years.” **KII-CFP**

The ease of installation of the ceiling net on the roof of traditional houses was an advantage and served as an indicator of appropriateness of fit of the intervention. The ceiling net took up the shape of the roof and does not affect any activities of daily living in the households. The respondents opined that the ceiling nets allowed for ventilation and less humid houses contributing to overall cleanliness and aesthetic appeal of the houses.

> “The design is well appropriate because it takes the shape of the roof, and this doesn’t bar the family or the household from doing any activities in the house. So, the way it was designed is very appropriate. There is no any complaint about it. In terms of lighting, ventilation and humidity, there is no problem that these people have experienced, because it is porous, the airflow is okay. In fact, instead it is more advantageous because now the flies and mosquitoes are being trapped from above, making the room to be very clean most of the time. In fact, they are adding even beauty to some houses.” **KII-SCPHO**

> **“**But in terms of, ah, maybe lighting and ventilation and humidity, there is no any… much problem that these people have experienced, because it is just a porous… the… the airflow is just okay. In fact, instead it is more advantageous because now the… the flies that are coming around, mosquitoes are being trapped from above, making the… the room to be very clean most of the time.” **KII-SCPHO**

Even as we report on the appropriateness of fit of the ceiling net as an innovative intervention of vector control, there is need for vigilance on resistance of the malaria parasites to insecticide compounds in the products.

> “If it gets used to one insecticide, we will not have anywhere to go because we will not have managed it. So can we have this combination or something else we can do apart from that.” **KII-SCP**

The design features of any intervention is a vital determinant of acceptability. The aesthetics in an intervention refers to a conscious consideration of the sensory and visual aspects to create a comfortable and friendly environment, in this case, malaria vector control. There was mention of the aesthetic appeal of the ceiling nets, adding beauty to the house.

> **“**The ceiling net is very much, very much appealing. In fact, they are adding even beauty to some houses.” **KII-SCPHO**

The health managers attributed the reduction in malaria in households using the ceiling nets to the ceiling nets’ efficacy.

> “Those who had the ceiling nets in their structures could appreciate that currently they don’t have several visits to the facility.” **KII-MoH**

There is need to demystify the myths and misconceptions regarding the ceiling nets.

> “There are myths and conceptions that we need to take away. Previously, somebody would report that I’m suffocating if I sleep under the bed net, which is not true. It is well aerated. The other thing, the ceiling net, the material that has been used does not prevent ventilation, and aeration will just be as usual.” **KII-MoH**

#### Sub-theme 2: Challenges with the use of ceiling nets

There is need to educate household members on how to clean dust and insects trapped on the nets, management of household pets such as cats that would tear through the ceiling nets in efforts of getting into the house through the eves.

> “They need to get it, from you or from us that once this has been provided, they should take care of the ceiling nets, not leaving the net so that it can be interfered with by the pets. They also need to know how to clean the insects trapped by the ceiling nets. They need to get this very clearly. That prompt in health education needs to be given so that they can use the best practices.” **KII-SCPHO**

> “They were saying the ceiling nets were very nice, but the pets that they have within their house, most can be the cats. If they cannot get access to the house, they have to get through the net, which means the nets will be torn so that the pets can get through.” **KII-MoH**

> “They were asking if I have a nice house with a ceiling net and now, I feel that it is now untidy or dirty or soiled, how can I take good care of it? Because with time, they get stained.” **KII-MoH**

#### Sub-theme 3: Suggested areas of ceiling net improvement

The hooks used to anchor the ceiling nets were mentioned as a potential area of improvement ensuring durability.

> “The main thing is we need to improve on the hooks. You know when you check on the durability of the hooks because you remember some could maybe hang every object in the room and that will interfere.” **KII-SCP**

Socio-behavioral change communication is imperative even as the community installs ceiling nets as a complementary vector control strategy. This would centre around ensuring that stagnant water is drained to eliminate mosquito breeding sites.

> “We need to continue telling them like the environmental control, because you find that in some places, yes you have the bed nets, but under the beds they could have dug some holes to hold water for them which are breeding sites for mosquito. So, if a behavior change, they could understand how this mosquito can be prevented from breeding and getting access to the house.” **KII-MoH**

## Discussion

This study demonstrates that health system managers perceive ceiling nets as a feasible, acceptable, and operationally integrable complement to existing malaria vector control strategies, while also identifying concrete system bottlenecks that must be addressed for scale-up. The World Health Organization highlighted the need for intersectoral collaboration for the implementation of home screening interventions such as ceiling nets [10]. Community health personnel play an instrumental role in the deployment and retrofitting of homes with the ceiling nets. This entails identification of housing structures with eaves, community mobilization, and installation of the ceiling nets. Open eaves are particularly relevant because they provide common mosquito entry points in many rural house types, making ceiling-level or eave-adjacent screening biologically plausible as a vector-control strategy. The current implementation model entails task-shifting where the community health promoters are trained in installation and maintenance of the ceiling nets, reducing the costs of implementation [15]. Health managers advocated for a complementary approach to large-scale deployment of insecticide treated nets and indoor residual spraying even with installation of ceiling nets, congruent with WHO guidelines on ceiling nets as a component of home screening.

A key barrier mentioned in the use of LLINs was the community concerns about discomfort, suffocation, or perceived adverse effects associated with sleeping under LLINs requiring a comprehensive community education and sensitization to improve LLIN’s programmatic effectiveness as propounded by Yirsaw et al. [16] in a study on barriers of persistent LLIN utilization in Ethiopia. The house arrangements in rural settings also limit the use of LLINs with indication of one bed net per bed usually for parents while children slept in the floor exposing them to mosquito bites. The ceiling net is highly effective in heterogeneous housing structures in rural Kenya marked by mud walls, thatched or iron roofs with open eaves, making them highly permeable to anopheles mosquitoes. The mosquitoes also tend to rest on ceilings where the nets can capitalize on this behavior effectively bringing them in contact with insecticide- treated nets. The installation of ceiling nets aims to extend protection beyond the sleeping spaces to the entire household [9]. The ceiling net is hailed as a house modification intervention that helps reduce mosquito entry while complementing the use of LLINs by extending the protective effect to other household members without access to LLINs [17]. In Peru, ceiling mounted mosquito nets resulted in a 55% reduction in count of indoor anopheles mosquitos and a 60% reduction in indoor bites per person per night [9]. The qualitative insights from the community focal person were in concurrence indicating a reduction in prevalence of malaria based on facility-based testing.

Community health promoters were hailed as critical community gatekeepers, instrumental in demand creation and raising awareness of malaria prevention efforts among community members. The gaps in their capacity development and remuneration were cited as imperative challenges in the integration of ceiling nets emphasized by reflections of health managers on the need for community knowledge transfer, a determinant of ceiling nets uptake and use. The presence of well-resourced, competent and trained community health personnel can contribute markedly to malaria control and elimination efforts through activities such as the distribution of insecticide-treated bed nets [18]. This study highlighted availability of commodity management systems as a fundamental health system facilitator that would be instrumental in ceiling net integration. Investing in a strengthened subnational public health supply chain is imperative to ensuring essential commodity availability thus reducing malaria-related morbidity and mortality [19].

The aesthetic appeal of the ceiling was mentioned as an important determinant of acceptability by the community with health managers indicating visual appeal as a facilitator of acceptability. This behavioral adoption dynamics, coupled with reduced user maintenance dependence and integradability with existing sleeping habits, makes the ceiling net an ideal complementary housing modification strategy in vector control. In accordance with these sentiments, eave screening with nets was found to make the house warmer and more attractive aesthetically whilst preventing malaria, in a study by Abong’o et al. [20] in rural Western Kenya. A key perceived challenge of the ceiling net in the study was the damage by house pets which was corroborated by a study that reported damage to house screens in 21% of the participants being caused by house animals [21].

The ceiling nets shows high acceptability, appropriateness of fit in structurally heterogenous housing structures and high feasibility as an addition to a comprehensive complementary LLIN and indoor residual spraying vector control strategy. Because ceiling nets may require less nightly user adherence than LLINs, health managers viewed them as a potentially sustainable complementary intervention. The ceiling net is a promising sustainable addition to the arsenal of vector control strategies [10], in rural areas of Kenya [8,22]. With recent studies indicating reduction in malaria parasites in children and decrease in indoor-resting mosquito density, the intervention shows efficacy in additional synergistic vector control in high endemic regions. The ongoing evaluation of cost-effectiveness will be critical in driving policy consideration in programmatic malaria prevention approach in Kenya.

This study is not without limitations that merit consideration. The perceptions reported herein were gathered from health managers requiring an in-depth look into the community perspectives regarding the ceiling nets. There is also need to correlate the findings herein on reduction in malaria prevalence with malaria surveillance data from health facilities. Having stated this, the findings provide generalizable perspectives on the health system readiness for integration of a complementary vector control strategy in malaria prevention.

## Conclusion

This study provides critical insights in enhancing integrated vector management in malaria endemic regions of Kenya. The findings inform decision-ready implementation pathways of complementary ceiling nets in existing vector control strategies. Critical barriers in the integration of ceiling nets in vector control strategies in a malaria endemic region were identified. These include underlying sociocultural perceptions of LLINs as the main complementary malaria and vector control intervention, lack of community health promoters’ capacity development and remuneration, and damage of ceiling nets by household pets. Key facilitators included available commodity management systems, extended protective effect and low behavioral dependency, and aesthetic appeal of the ceiling nets. The ceiling net, in particular the insecticide-treated Olyset®Plus ceiling net, is well positioned in Kenya’s Integrated vector management policy as a promising complementary tool. We show that ceiling nets are perceived by frontline health managers as a system-ready, acceptable house-modification intervention that can close critical protection gaps left by bed-net–centric malaria control, provided community health platforms and supply chains are deliberately leveraged. The findings underpin the importance of continuous iterative learning as we design contextually responsive tools in the next generation of malaria vector control. Successful scale up implementation hinges on a structured approach of integration that includes community buy-in, task-shifting of community health promoters in ceiling net installation, effective supply chains, and robust monitoring and evaluation systems to track, maintain and assess system level indicators and community level indicators.

## Data Availability

The qualitative data, including the de-identified interview transcript excerpts relevant to this study, are publicly available and deposited in the Open Science Framework repository under the following Link:https://osf.io/wpu7m/overview.

## Acknowledgements

We wish to thank all the sub-county health system managers who contributed their valuable insights to this study on health systems readiness. We also acknowledge the institutional support provided by Mount Kenya University and Homa Bay County Department of Health.

